# Specific allelic discrimination of N501Y and other SARS-CoV-2 mutations by ddPCR detects B.1.1.7 lineage in Washington State

**DOI:** 10.1101/2021.03.10.21253321

**Authors:** Garrett A. Perchetti, Haiying Zhu, Margaret G. Mills, Lasata Shrestha, Cassia Wagner, Shah Mohamed Bakhash, Michelle Lin, Hong Xie, Meei-Li Huang, Patrick Mathias, Trevor Bedford, Keith R. Jerome, Alexander L. Greninger, Pavitra Roychoudhury

## Abstract

Real-time epidemiological tracking of variants of interest can help limit the spread of more contagious forms of SARS-CoV-2, such as those containing the N501Y mutation. Typically, genetic sequencing is required to be able to track variants of interest in real-time. However, sequencing can take time and may not be accessible in all laboratories. Genotyping by RT-ddPCR offers an alternative to sequencing to rapidly detect variants of concern through discrimination of specific mutations such as N501Y that is associated with increased transmissibility. Here we describe the first cases of the B.1.1.7 lineage of SARS-CoV-2 detected in Washington State by using a combination of RT-PCR, RT-ddPCR, and next-generation sequencing. We screened 1,035 samples positive for SARS-CoV-2 by our CDC-based laboratory developed assay using ThermoFisher’s multiplex RT-PCR COVID-19 assay over four weeks from late December 2020 to early January 2021. S gene dropout candidates were subsequently assayed by RT-ddPCR to confirm four mutations within the S gene associated with the B.1.1.7 lineage: a deletion at amino acid (AA) 69-70 (ACATGT), deletion at AA 145, (TTA), N501Y mutation (TAT), and S982A mutation (GCA). All four targets were detected in two specimens, and follow-up sequencing revealed a total of 10 mutations in the S gene and phylogenetic clustering within the B.1.1.7 lineage. As variants of concern become increasingly prevalent, molecular diagnostic tools like RT-ddPCR can be utilized to quickly, accurately, and sensitively distinguish more contagious lineages of SARS-CoV-2.

## BACKGROUND

The first known case of the SARS-CoV-2 B.1.1.7 variant in the United States was reported in Colorado on December 29^th^ 2020 and the next day, it was confirmed in California (1, 2). University of Washington (UW) Virology had begun surveillance a few days prior using PCR to screen SARS-CoV-2 positive samples for the absence, or “dropout” of the S gene, which encodes the spike (S) protein on the surface of the viral particle. The B.1.1.7 variant is characterized by 17 mutations, eight of which occur within the S gene domain (3, 4). This region of the SARS-CoV-2 genome is of interest due to the B.1.1.7 lineage being associated with increased transmissibility, but also because the FDA emergency use authorization for COVID-19 vaccines in the United States target the S protein (5, 6).

Over the course of four weeks, we screened more than a thousand SARS-CoV-2 positive samples for the S gene target failure (SGTF). We selected random clinical specimens that were positive using a CDC-based laboratory developed test (LDT) for SARS-CoV-2, and typically had a cycle threshold (C_T_) under 35 (7–11). These SARS-CoV-2 positive samples were then amplified with the TaqPath COVID-19 assay (ThermoFisher Scientific, Waltham, MA, USA), a multiplex RT-PCR targeting the S gene, as well as regions within the N gene and ORF1ab (12). Candidates for the B.1.1.7 variant have a positive detection for the N gene and ORF1ab, with a negative for the S gene. SGTFs are candidates for the B.1.1.7 lineage, but the TaqPath assay is not necessarily specific for that variant exclusively due to other signature mutations, so genetic sequencing is used to confirm the B.1.1.7. lineage (13). However, sequencing can be a time-consuming and resource-intensive process that not all laboratories have integrated into their clinical workflow. Analysis of publicly available sequencing data by the Broad Institute revealed that it takes a median 85 days to get from sample to publicly available sequence in the United States (14, 15).

Here we describe a novel droplet reverse-transcription digital-PCR (RT-ddPCR) assay that specifically detects four mutations associated with the B.1.1.7 variant, particularly the N501Y mutation. This mutation in the S gene is shared by the U.K. and the South African variant (B.1.351) (16). Preliminary data has indicated B.1.1.7 to be more transmissible, while B.1.351 is considered to be less well neutralized by antibodies induced by certain vaccines, such as the AstraZeneca and Novavax vaccines (3, 16–18). This RT-ddPCR assay can distinguish SARS-CoV-2 positive samples that carry this important N501Y mutation, as well specific allelic discrimination of the B.1.1.7 lineage, without genetic sequencing.

## METHODS

### Specimen Selection Criteria

From December 25^th^ 2020 to January 20^th^ 2021, 1,035 specimens positive for SARS-CoV-2 by our CDC-based LDT were screened for SGTFs (Fig.1) (19–21). Approximately 50% of these samples came from King County, followed by Pierce County with around 15%, Benton and Franklin Counties at 10%, and the remainder from other counties in Washington State. We selected samples with C_T_s<35 when available to reduce the effect of assay stochasticity, with consideration of initial data showing that the B.1.1.7 variant has been associated with higher viral loads (i.e. lower C_T_s) (22). Extracted nucleic acids were stored at 4°C or -20°C prior to amplification. This work was approved under a waiver of consent by the University of Washington institutional review board (STUDY00000408).

**Figure 1.**
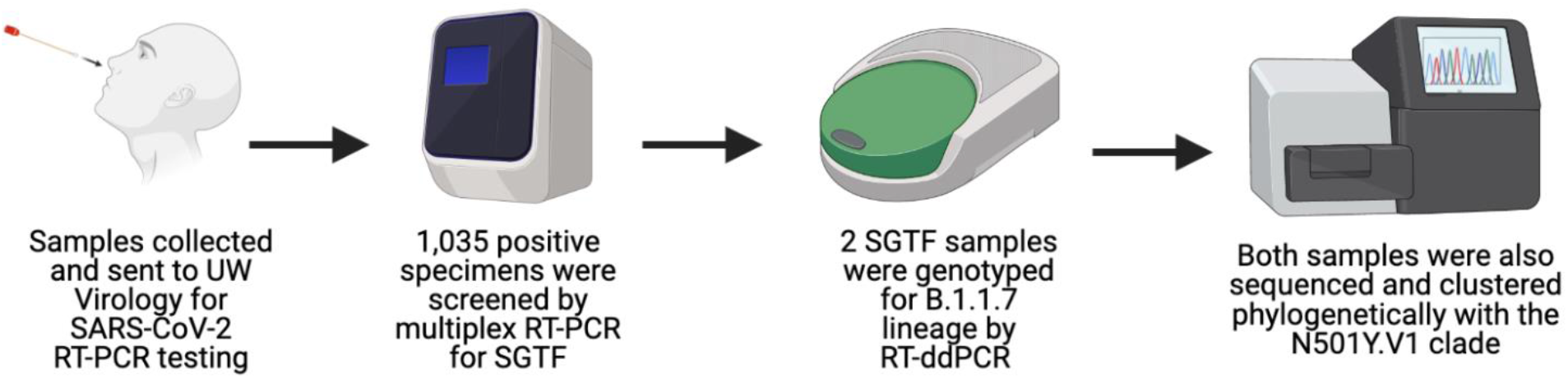
RT-PCR, RT-ddPCR, and sequencing surveillance effort for detection of B.1.1.7 lineage Samples sent to UW Virology for SARS-CoV-2 molecular detection were screened for S gene transcript failure (SGTF) by multiplex RT-PCR as a proxy for variant detection. SARS-CoV-2 positive samples with SGTF were subsequently genotyped for specific allelic discrimination by RT-ddPCR. Both SGTF samples that genotyped as B.1.1.7 lineage by RT-ddPCR were also sequenced and clustered with N501Y.V1 clades in the United States.

### PCR

PCR was performed using TaqPath COVID-19 Combo Kit (ThermoFisher, Waltham, MA, USA) with 10 µL of extracted nucleic acid used as template per 25µL reaction. This multiplex real-time RT-PCR assay targets the S gene, N gene, and ORF1ab of SARS-CoV-2. Reactions utilized a kit-provided positive control (1×10^4^ copies/µL) diluted with TaqPath COVID-19 Control Dilution Buffer, and dH_2_O as a negative template control. Amplifications were run on Applied Biosystems 7500 Real-Time PCR Systems (ThermoFisher) according to manufacturer’s thermocycling parameters.

### RT-ddPCR

Four sets of primers and probes were designed based on U.K. variant sequence hCoV-19/England/MILK-9E2FE0/2020 (EPI_ISL_581117, collection date 2020-09-21). To check for variation at primer and probe sites, 216 B.1.1.7 sequences were downloaded from GISAID on December 23^rd^, 2020, and aligned against the Wuhan-Hu1 reference sequence (NC_045512.2) using MAFFT v7.450 within Geneious Prime (https://www.geneious.com) (Supplementary Table 1). In addition, a G-block of 490bp was designed that includes four target amplicons. All primer and probes were synthesized by ThermoFisher and the G-block was synthesized by IDT (Coralville, IA, USA). Primers were included at 900 nM and probes were used at 250 nM concentrations. All four targets are within the S gene domain: a deletion at amino acid (AA) 69-70 (ACATGT), deletion at AA 145, (TTA), N501Y mutation (TAT), and S982A mutation (GCA). The specific primers and probe sequences and characteristics are outline in Table 1; the G-block sequence: AACTCAGGACTTGTTCTTACCTTTCTTTTCCAATGTTACTTGGTTCCATGCTATCTCTGGGACCAATGGTACTAAGAGGTTTGATAACCCTGTCCTACCATTTAATGATGACGCTACTAATGTTGTTATTAAAGTCTGTGAATTTCAATTTTGTAATGATCCATTTTTGGGTGTTTACCACAAAAACAACAAAAGTTGGATGGAAAGTGAGTTCAGAGTTTATTCTAGTGCGAATAATTGTACACACCTTGTAATGGTGTTGAAGGTTTTAATTGTTACTTTCCTTTACAATCATATGGTTTCCAACCCACTTATGGTGTTGGTTACCAACCATACAGAGTAGT AGTACTTTCTTTTGAACTTCTACATGCACCAGCAACCCAAACAACTTAGCTCCAATTTTGGTGCAATTTCAAGTGTTTTAAATGATATCCTTGCACGTCTTGACAAAGTTGAGGCTGAAGTGCAAATTGATAGGTTGATCACAGGC.

**Table 1.**
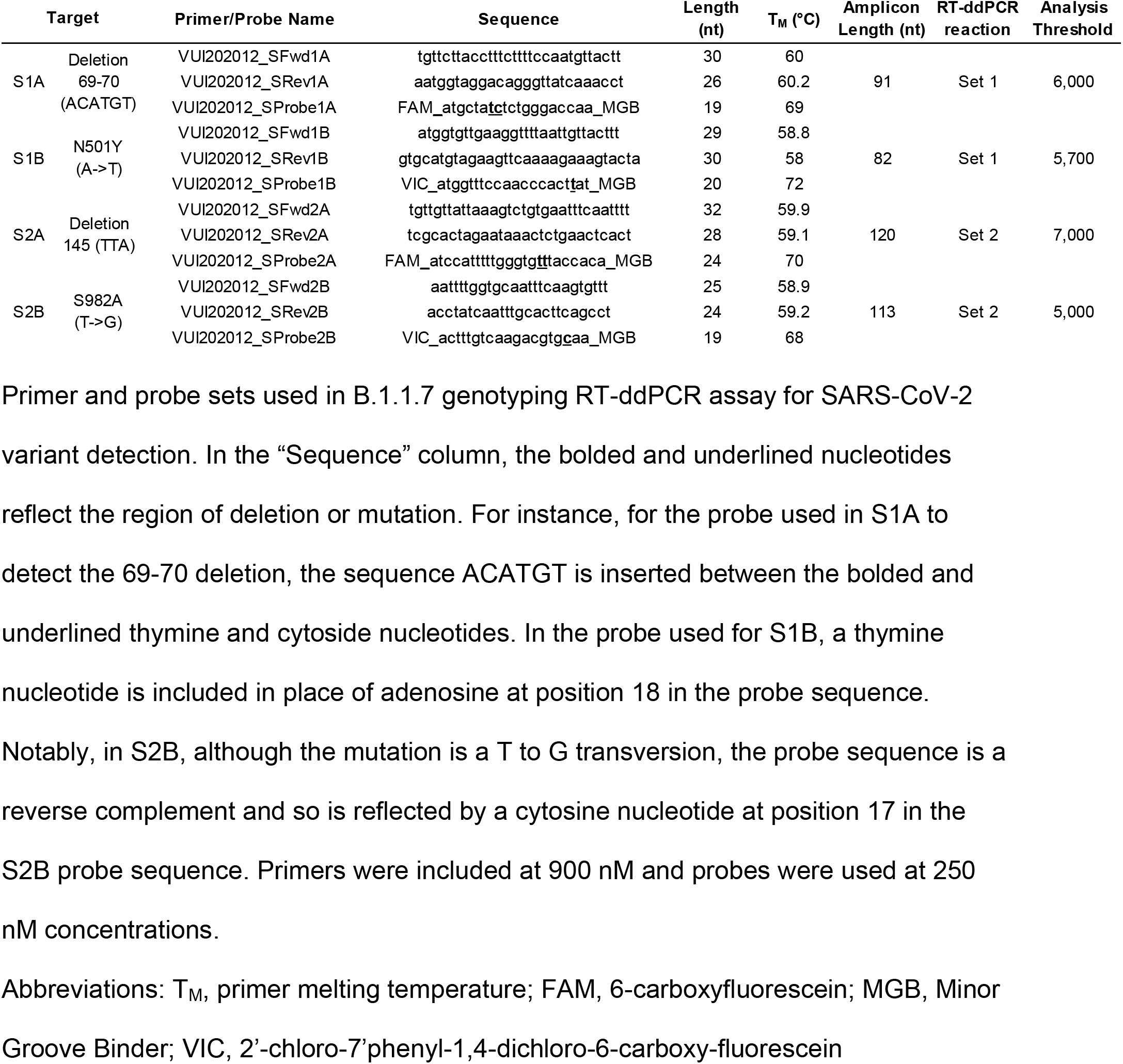
Design information for B.1.1.7 identification assay.

Two multiplex RT-ddPCR reactions per sample were performed in parallel (as outlined in Table 1) using One-step RT-ddPCR Advanced Kit for Probes (Bio-Rad Laboratories, Hercules, CA, USA) with the Automated Droplet Generator (Bio-Rad) and C1000 Touch thermocycler (Bio-Rad). Template RNA for each clinical sample was diluted to an approximate N1 C_T_ of 30 before amplification. Thermocycling conditions were as follows: 50°C for 60 min, 40 cycles at 95°C for 30 sec and 60°C for 1 min, then 98°C for 10 min. Droplet detection was performed using the QX200 Droplet Reader (Bio-Rad) and QuantaSoft Pro 1.0.596 version software. For SARS-CoV-2 B.1.1.7 detection, all four targets are amplified beyond the thresholds. For non-B.1.1.7 samples, 0-1 target(s) are amplified beyond the thresholds. Both the synthetic G-block and a known B.1.1.7 clinical sample are used as positive controls, and a SARS-CoV-2 clinical positive control is used as a negative control in the B.1.1.7 assay.

### Sequencing

For next-generation sequencing, 11 µL of extracted RNA was used for single-stranded complementary DNA (sscDNA) synthesized using SuperScript IV First-Strand Synthesis System according to the manufacturer’s protocol (ThermoFisher). Libraries were prepared using the Swift Normalase Amplicon Panel (SNAP) for SARS-CoV-2 (Swift Biosciences, Ann Arbor, MI, USA) as previously described (23). For standards, a wild type clinical nasopharyngeal specimen positive for SARS-CoV-2 was used as a positive control and water was used as no-template negative control. The resulting libraries were quantified fluorometrically on Qubit 3.0 using the Quant-iT dsDNA high sensitivity kit (Life Technologies, Carlsbad, CA, USA). The libraries passing quality control, nucleic acid concentrations > [1.3 ng/µL], were normalized manually and sequenced on the Illumina MiSeq platform (Illumina, San Diego, CA, USA) using MiSeq Reagent Kit v2 (2×150 reads).

Raw reads were analyzed using a custom bioinformatic pipeline (TAYLOR, https://github.com/greninger-lab/covid_swift_pipeline) (23). Briefly, raw reads were trimmed to remove adapters and low-quality regions and mapped to the Wuhan-Hu-1 reference sequence (NC_045512.2) using BBMap v38.86 (https:/jgi.doe.gov/data-and-tools/bbtools/). Aligned reads were then soft-clipped of PCR primers using the PrimerClip package from Swift Biosciences (https://github.com/swiftbiosciences/primerclip) and a consensus sequence was called using bcftools v1.9 (24). Consensus sequences were aligned using MAFFT v7.450 within Geneious Prime (https://www.geneious.com) (25). Mutations in the spike protein were manually reviewed in addition to automated variant calling within the pipeline. Clade assignment was performed using Nextclade v0.12.0 (https://github.com/nextstrain/nextclade). Sequences were deposited to Genbank (accessions pending) and GISAID (EPI_ISL_861730 and EPI_ISL_861731); raw reads were deposited to the NIH’s Sequence Read Archive (Bioproject PRJNA610428).

Phylogenetic tree construction utilized the Nextstrain pipeline to align sequences, reconstruct maximum-likelihood and time-resolved phylogenetic trees and to infer nucleotide and amino acid substitutions across the phylogeny (26). The specific workflow for this analysis is available at: https://github.com/blab/ncov-wa-build. The tree included the B.1.1.7 sequences described here, 1,586 SARS-CoV-2 samples available on GISAID collected in Washington State from November 2020 through February 12^th^, 2021, and an additional 1,824 global contextual sequences from GISAID sampled based on genetic similarity to the Washington sequences (Supplementary Table 2). The tree, continually updated with additional Washington sequences collected between November 2020 and February 2021, can be viewed at: https://nextstrain.org/groups/blab/ncov/wa/nov20-feb21.

## RESULTS

### PCR

Means, medians, and the ranges of C_T_s detected for the S gene, N gene, and ORF1ab targets are characterized in Table 2. Seven samples were candidates for SGTF, out of a total of 1,035 samples screened for the B.1.1.7 variant. Of these, 5/7 had C_T_s >33.5 for all targets with a mean and median C_T_ of 35.7 and 35.6 for N gene, and C_T_s 37.8 and 37.5 for ORF1ab, respectively. Two samples however, had strong fluorescent amplification for the N gene and ORF1ab targets, but no S gene amplification (Fig. 2). Candidate 1 (55538) had C_T_s of 23.4 and 23.7 for N gene and ORF1ab targets respectively, without S gene amplification. Candidate 2 (55545) had C_T_s of 26.7 and 26.4 for N gene and ORF1ab. Consequently, we decided to ddPCR and sequence these specimens.

**Table 2.** TaqPath COVID-19 assay cycle thresholds characteristics of SARS-CoV-2 specific targets screening for S gene dropouts.

| Target | C<sub>T</sub> mean | C<sub>T</sub> median | C<sub>T</sub> range | No. Detected |
| --- | --- | --- | --- | --- |
| N gene | 24.2 | 22.9 | 14.4-38.9 | 1,035 |
| ORF1ab | 23.5 | 22.3 | 14.3-39.8 | 1,001 |
| S gene | 24.1 | 22.9 | 14.5-39.8 | 994 |

Three SARS-CoV-2 specific amplicons, N gene, ORF1ab, and S gene, were targeted with the multiplex RT-PCR ThermoFisher COVID-19 Assay. Over 1,000 samples positive for SARS-CoV-2 were screened for SGTF, with seven candidates having N gene and ORF1ab amplification without S gene detection. Two of these seven candidates were determined to candidates for genotyping by RT-ddPCR and sequencing based on their viral load characteristics (i.e. mean and median C<sub>T</sub>S> 35.0 for all targets detected).
Abbreviations: C<sub>T</sub>, cycle threshold, SGTF, S gene target failure

**Figure 2.**
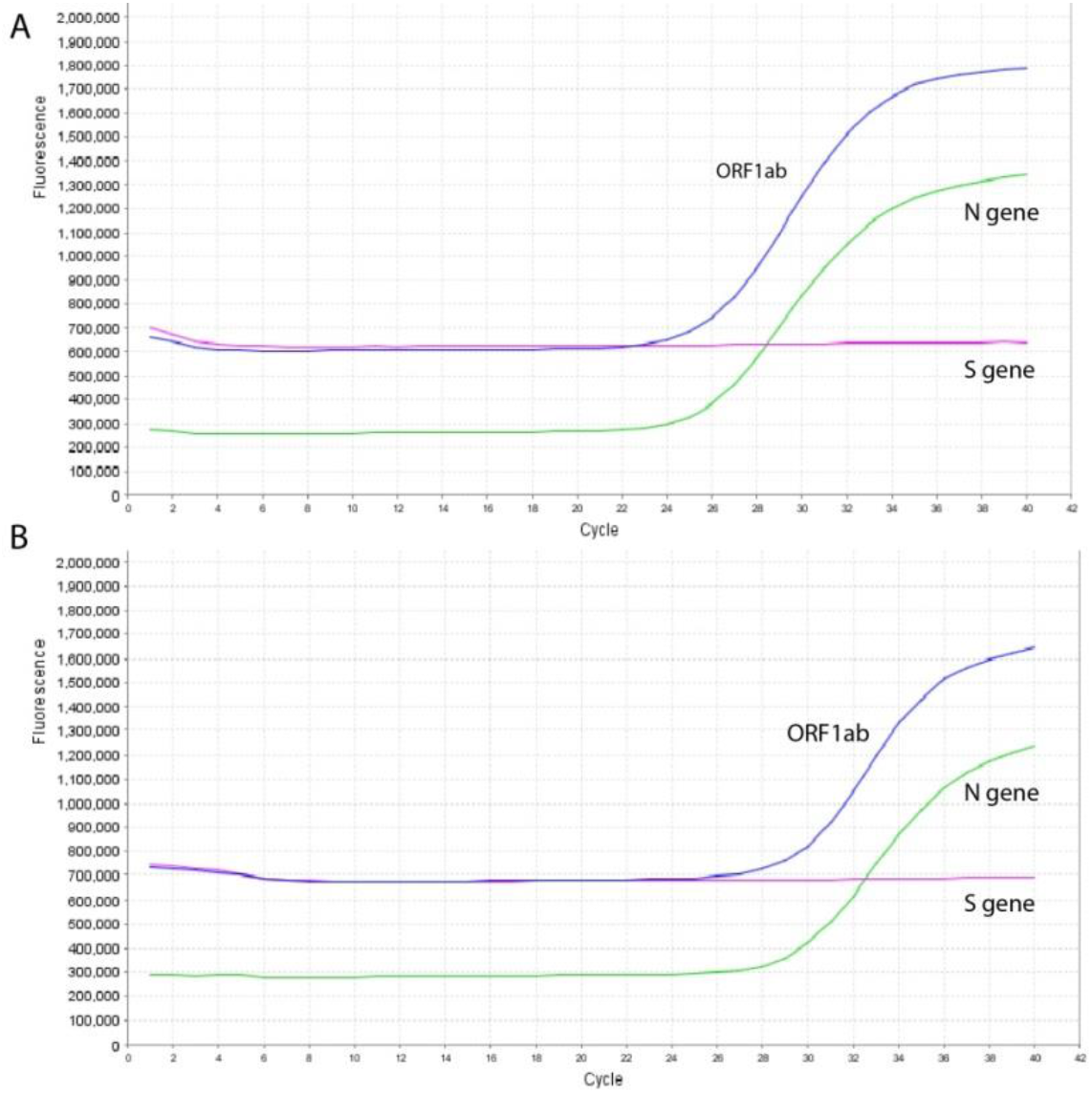
Multiplexed qRT-PCR fluorescence of S gene dropout in SARS-CoV-2 positive samples Here, multicomponent amplification plots are shown for (A) U.K. Variant #1 (55538) and (B) U.K. Variant #2 (55545). PCR cycle is plotted on the X axis, with quantity of fluorescence detected in real-time on the Y axis. Two out of three fluorophores are detected by qRT-PCR for both samples on the 7500 Real-Time PCR System (Life Technologies, Carlsbad, CA, USA) and analyzed on 7500 software v.2.3 (Life Technologies). VIC and FAM are reporters for N gene and ORF1ab, respectively and are seen here with robust amplification. ABY, the probe for the S gene, is present in the reaction mix, but is not detected by fluorescence, indicating a potential candidate for a SARS-CoV-2 variant

### RT-ddPCR

Both specimens showed clear fluorescent amplification above analysis thresholds of all four targets for both sets of RT-ddPCR reactions (Fig. 3). Amplification characteristics are outlined in Table 3 with absolute virus copies/µL of RNA included in the ddPCR reaction. With back-calculations considering RNA dilutions and extraction compressions, U.K. Variant #1 (55538) was quantified to have 765,500-860,000 virus copies/mL and U.K. Variant #2 (55545) was quantified to have 60,250-72,250 virus copies/mL depending on target amplicon.

**Table 3.** RT-ddPCR genotyping results for two SGTF samples.

| Sample | S1A | S1B | S2A | S2B |
| --- | --- | --- | --- | --- |
| U.K. Variant #1_55538 | 344 (860,000) | 306 (765,000) | 323 (807,500) | 336 (840,000) |
| U.K. Variant #2_55545 | 289 (72,250) | 241 (60,250) | 267 (66,750) | 275 (68,750) |
| Pos Control<sup>1</sup> | 705 | 680 | 588 | 630 |
| Pos Control<sup>2</sup> | 660 | 733 | 654 | 833 |
| Neg Control<sup>1</sup> | 0 | 0 | 0 | 0 |
| Neg Control<sup>2</sup> | 0 | 0 | 0 | 0 |

Copies per µl RNA for each target amplified in multiplex RT-ddPCR reaction.
Parentheticals denote virus copies/mL back-calculated to consider RNA extraction and
RT-ddPCR dilution factors. Pos control = <sup>1</sup>Previously-identified and sequenced B.1.1.7 clinical sample; <sup>2</sup>G-block. Neg control = <sup>1</sup>Clinical (non-B.1.1.7) SARS-CoV-2 positive; <sup>2</sup>water.
Abbreviations: SGTF, S gene target failure

**Figure 3.**
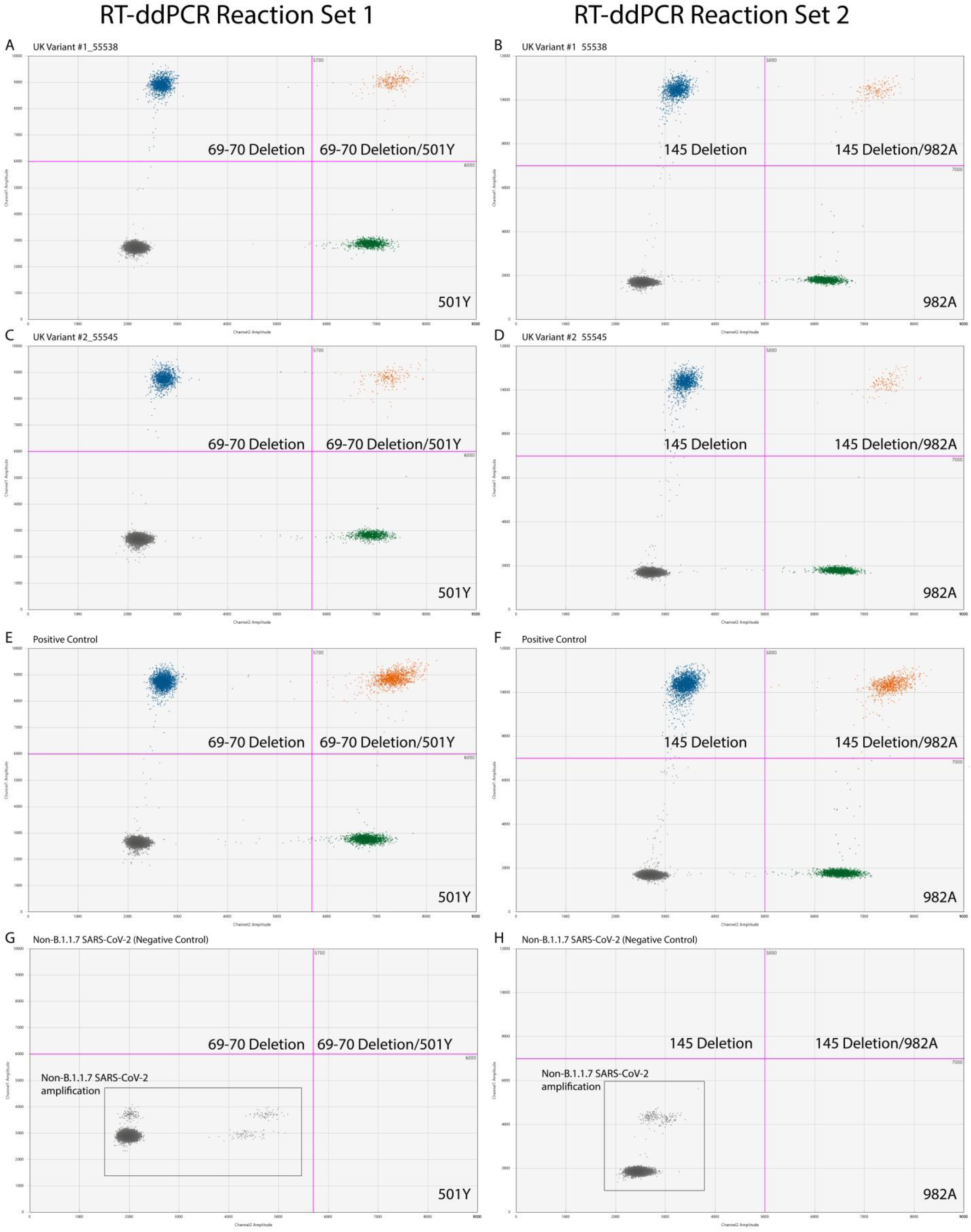
RT-ddPCR amplification results for SARS-CoV-2 B.1.1.7 lineage (A) U.K. Variant Sample #1 (55538) and (C) U.K. Variant Sample #2 (55545) have amplification for AA69-70del and N501Y mutation. (E) Positive control demonstrates amplification for AA69-70del and N501Y mutation and (G) B.1.1.7 Negative control (Non-B.1.1.7 SARS-CoV-2) shows no amplification for AA69-70del and N501Y mutation for RT-ddPCR amplicon set 1. (B) U.K. Variant Sample #1 (55538) and (D) U.K. Variant Sample #2 (55545) have amplification for AA145 deletion and 982A mutation. (F) Positive control demonstrates amplification for AA145 deletion and 982A mutation and (G) B.1.1.7 Negative control (Non-B.1.1.7 SARS-CoV-2) shows no amplification for AA145 deletion and 982A mutation for RT-ddPCR amplicon set 2.

While the primer sites for all four targets are present in wild type SARS-CoV-2 viral RNA as well as B.1.1.7 RNA, the probes for each target are designed to bind specifically to the regions mutated in the B.1.1.7 variant. Depending on the difference in melting temperature between the wild type and variant sequences for a given target, the probe may show some binding to the wild type sequence as well, but at a lower efficiency than it binds to the variant sequence. In RT-PCR, inefficient binding to the wild type strand is indistinguishable from high-efficiency binding to a lower-concentration variant strand. In RT-ddPCR, because individual template strands are amplified in separate droplets, inefficient probe binding can be identified as lower-amplitude fluorescence from each droplet. Thus, even an A to T single nucleotide polymorphism such as that present in the N501Y mutation (S1B) is easily distinguishable by RT-ddPCR by screening for droplets with S1B probe amplitude above a threshold of 5,700.

### Sequencing

We obtained good quality consensus genomes for both samples, with more than 20,000X average coverage across the genome (Table 4). Both samples were classified as 20I/501Y.V1 using Nextclade (ver 0.12.0) and had 100% pairwise nucleotide identity in a whole genome alignment). For each sample, a total of 10 mutations were found in the spike protein: H69-, V70-, Y144-, N501Y, A570D, D614G, P681H, T716I, S982A, D1118H. In a phylogenetic tree, these samples represented a unique cluster within clade 20I/501Y.V1 (Figure 4). They are 6 mutations diverged from the genetically closest sample available in GISAID, England/ALDP-BB47ED/2020, sampled in the United Kingdom in November 2020. Other B.1.1.7 samples collected in Washington did not cluster with them, suggesting that these samples represent a unique introduction into the state.

**Table 4.**
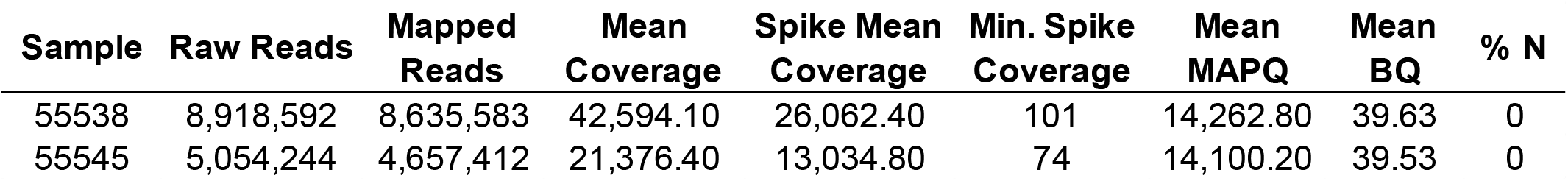
Next-generation sequencing characteristics.

| Sample | Raw Reads | Mapped Reads | Mean Coverage | Spike Mean Coverage | Min. Spike Coverage | Mean MAPQ | Mean BQ | % N |
| --- | --- | --- | --- | --- | --- | --- | --- | --- |
| 55538 | 8,918,592 | 8,635,583 | 42,594.10 | 26,062.40 | 101 | 14,262.80 | 39.63 | 0 |
| 55545 | 5,054,244 | 4,657,412 | 21,376.40 | 13,034.80 | 74 | 14,100.20 | 39.53 | 0 |

**Figure 4.**
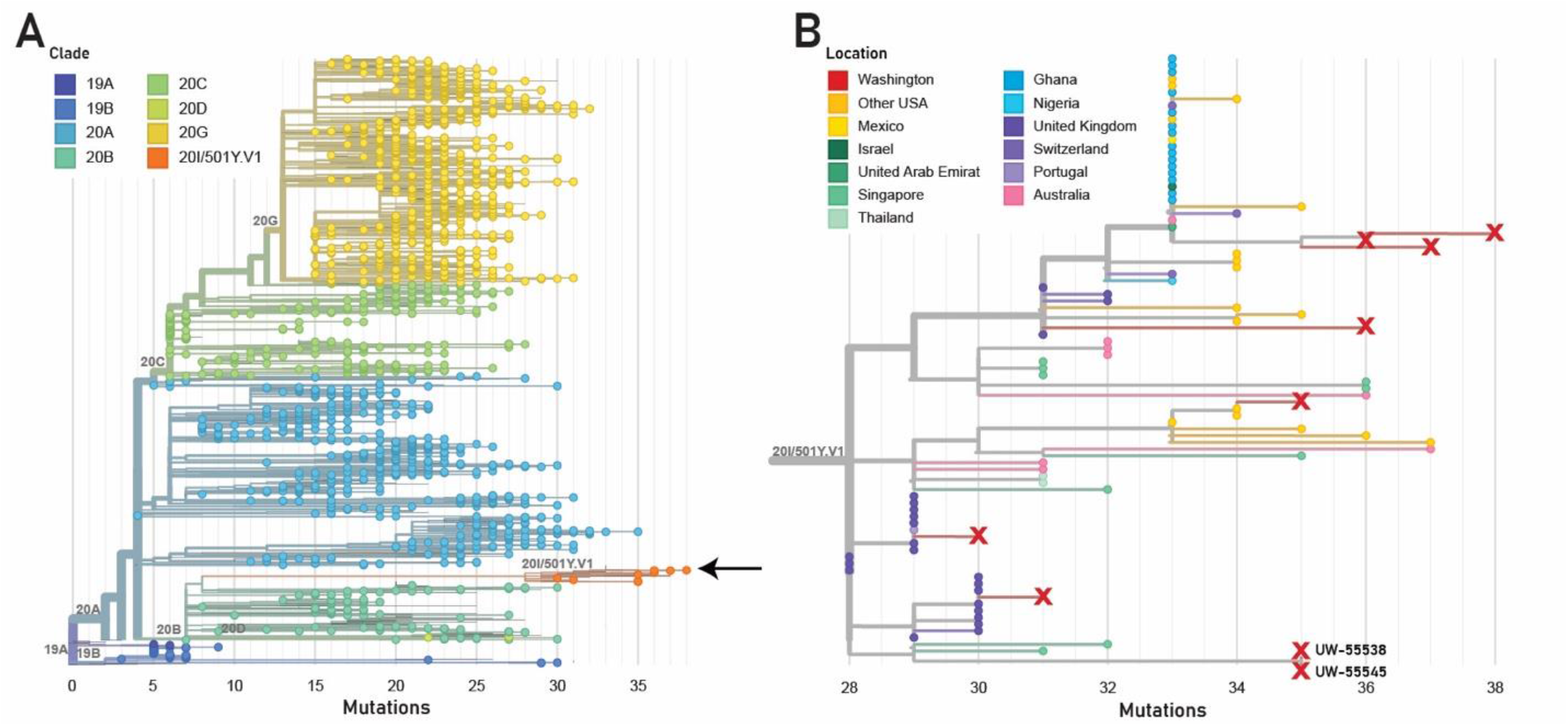
Phylogenetic tree focused on Washington SARS-CoV-2 samples collected from November 2020 through February 2021 In (A) the phylogenetic tree is filtered to only show Washington samples; 501Y.V1 (B.1.1.7) samples are shown in orange (arrow). (B) 501Y.V2 clade showing Washington samples (red X) in context of global SARS-CoV-2, selected by genetic proximity to the Washington samples. UW-55538 &UW-55545 separately cluster at the bottom of the clade.

## DISCUSSION

The B.1.1.7 variant of SARS-CoV-2 is more transmissible than the original lineage, which can ultimately lead to an increase in global cases, unfortunately resulting in more death. Detection of this variant has clinical and epidemiological relevance due to its increased transmissibility, as well as the N501Y mutation being shared with the South African (B.1.351) variant (27). B.1.351 is considered to be associated with decreased immunogenicity by provoking a weaker neutralizing antibody response against spike protein mutants (18, 28). Unfortunately, this means recently developed vaccines may have diminished vaccine efficacy. Pfizer reported that via plaque reduction neutralization testing, neutralization of the B.1.351 variant was reduced by two-thirds compared to USA-WA1/2020 (29); in the same day, Moderna reported their vaccine elicited sixfold less antibodies against the B.1.351 variant (30). These mutations have tremendous real-world impact as South Africa recently halted using the Oxford-AstraZeneca vaccine due to its decreased efficacy against the B.1.351 variant (31).

Our experience with the SGTF screening underscores the enduring utility of multiplex PCR assays and their adaptability. PCR remains a powerful tool in the scalability of screening large volumes of viral samples for variant detection. Other clinical laboratories from San Francisco to Lyon have also recently implemented modified PCR-based screening methods to hone in on samples with potential to be variants of interest (32–34). Using PCR and allelic discrimination ddPCR in conjunction with sequencing, clinical laboratories can efficiently and quickly identify SGTF specimens that are candidate variants of concern. Helix reported that of the positive COVID-19 tests screened, less than 1% had SGTF, however, of those SGTFs, more than one-third were confirmed B.1.1.7 lineages (35).

According to GISAID, the United States has only sequenced 3 out of every 1,000 positive SARS-CoV-2 samples (36). RT-ddPCR has potential to identify samples to prioritize for sequencing, allowing more efficient allocation of strain surveillance resources. However, multiple targets are necessary to accurately detect newer viral mutations, such as those seen in the U.K. and South African variants; these mutations can arise in regions that are targeted by PCR primers (37, 38). Assays that only target the S gene domain of SARS-CoV-2 run the risk of missing newer variants and ultimately lead to an increase in false negatives (39).

Although our surveillance was limited, the variant positivity at time of initial detection was approximately 1 in 500 (2 out of 1,035) randomly selected SARS-CoV-2 samples. However, reports indicate that due to B.1.1.7’s increased transmissibility, it is already increasing in frequency and is soon expected to become the dominant strain in the United States (6, 27, 35, 40). The large number of genomic mutations associated with B.1.1.7, including single nucleotide polymorphisms (SNPs) A570D, D614G, P681H, T716I, S982A, and D1118H, underscores the need to distinguish these subtle mutations in emerging variants of concern (41). RT-ddPCR is more sensitive than RT-PCR at resolving SNPs, and is better suited for allelic discrimination to differentiate the B.1.1.7 lineage or other variants of concern (42–44). The S982A SNP, for instance, is specific for B.1.1.7 lineages, whereas the N501Y mutation is not (40). The S982A SNP is readily detectable using RT-ddPCR, however, would not be distinguishable by RT-PCR.

As genetic surveillance becomes increasingly relevant in efforts to track and understand new SARS-CoV-2 variants of concern in real-time, RT-ddPCR continues to cement its place in the clinical laboratory armamentarium (15, 17, 26, 41, 45). However, RT-ddPCR technology is still not ubiquitous in clinical laboratory settings. Increased adoption and investment in this technology can allow labs to rapidly estimate prevalence of existing variants and perform sample screening to better allocate limited sequencing resources.

## Data Availability

Data has been upload to GISAID, Genbank, and SRA, and code is available via Github.

## AKNOWLEDGEMENTS

We gratefully acknowledge the authors and laboratories involved in the generation and deposition of sequencing data, which we obtained via the GISAID Initiative (Supplementary Table 2). The authors would also like to thank Victoria Mallett Rachleff for assistance in data deposition.

